# Diagnostic accuracy of LRINEC score and vitals in Necrotising skin infection in clinical Surgical Scar Incision Infection: A meta-analysis

**DOI:** 10.1101/2023.06.16.23291501

**Authors:** Marlene Carina Götz, Dev Andharia, Maria Eleni Malafi, Pankti Maniyar, Dwija Raval, Siddharth Agrawal, Dev Desai

## Abstract

**Background:** Necrotizing soft tissue infection (NSTI) is a potentially fatal skin and soft tissue infection, characterized by fulminant tissue damage, systemic signs of toxicity, and high mortality with case fatality rates ranging from 14% to 41% over the last two decades. It can be challenging to diagnose NF in its initial phases as it frequently presents symptoms that are similar to those of other non-necrotic SSTIs, such as cellulitis. It is unclear how the different diagnostic imaging modalities should be used to evaluate patients who have a suspected NSTI and there are concerns about their accuracy and potential delays in surgical intervention. Therefore, we aimed to gather data on the sensitivity and specificity of physical findings of fever, hypotension as well as imaging techniques such as ultrasound (USG) and computed tomography (CT) scans, and the LRINEC score, in detecting Necrotizing Soft Tissue Infections (NSTI) in patients.

**Methods:** Medical literature was comprehensively searched and reviewed without restrictions to particular study designs, or publication dates using PubMed, Cochrane Library, and Google Scholar databases for all relevant literature. The extraction of necessary data proceeded after specific inclusion and exclusion criteria were applied. In this Meta-Analysis, a total of 49 RCTs with an aggregate of 11,520 cases were handpicked. wherein two writers independently assessed the caliber of each study as well as the use of the Cochrane tool for bias risk apprehension. The statistical software packages RevMan (Review Manager, version 5.3), SPSS (Statistical Package for the Social Sciences, version 20), and Excel in Stata 14 were used to perform the statistical analyses.

**Results:** We calculated the sensitivity and specificity for each of the parameters. Here, USG has a sensitivity of 0.556 and specificity of 0.879, CT has a sensitivity of 0.932 and specificity of 0.849, and LRINEC Score >= 6 has a sensitivity of 0.59 and specificity of 0.849. we also calculated the same for physical signs like fever and hypotension.

**Conclusion:** we conclude that physical signs like fever and hypotension and LRINEC Score >= 6 are not advisable indicators, however, CT shows significant superior modality but it is not a cost-effective solution. USG is relatively reliable and cost-effective for the early diagnosis of NSTI.

## INTRODUCTION

Necrotizing soft tissue infection (NSTI) is a potentially fatal skin and soft tissue infection, characterized by fulminant tissue damage, systemic signs of toxicity[1], [2]and high mortality with case fatality rates ranging from 14% to 41% over the last two decades.[3], [4],[5] [6], [7], [8],[9]. It can be challenging to diagnose NF in its initial phases as it frequently presents symptoms that are similar to those of other non-necrotic SSTIs, such as cellulitis[10], [11]. Doctors typically rely on physical examination, radiologic imaging methods such as USG, CT, and MRI, and clinical decision instruments to diagnose NSTI, but there is little evidence to support the usefulness of these diagnostic tools.Signs like severe pain which is out of proportion to physical findings, fever, crepitus, and necrosis are used clinically, to differentiate NSTI from soft tissue infections. Systemic findings like hypotension and shock are also more suggestive of NSTI [12][13][14]. Radiologic imaging tools like X-ray, USG, MRI, and CT have been used to aid the diagnosis. CT is considered as best initial imaging test but is usually time-consuming and can cause delays in surgical management.[15], [16]. The presence of gas in soft tissue is highly specific for NSTI but may be restricted to anaerobic bacterial infections or advanced disease phases[17]. Bedside USG, being rapid and easily available, is commonly used in emergency departments for the detection of soft tissue skin infections but is not very well studied for use in identifying NSTI[18]. Lab findings are usually non-specific, but a tool called the Laboratory Risk Indicator for Necrotising Fasciitis (LRINEC) score is sometimes used to predict the chances of NSTI. It is based on the routinely performed laboratory tests: C-reactive protein (CRP), white cell count, hemoglobin, serum sodium, serum creatinine, and glucose levels. An LRINEC score of 6 should raise the suspicion of necrotizing fasciitis, and a score of 8 is strongly predictive of this disease.[19], [20].It is unclear how the different diagnostic imaging modalities should be used to evaluate patients who have a suspected NSTI and there are concerns about their accuracy and potential delays in surgical intervention. These investigations may help prevent unnecessary operations and assist in the planning of the operative exploration[21], [22]. Due to the prevalence of conflicting schools of thought and the limited sample sizes of many studies, our primary objective in conducting a comprehensive review and meta-analysis was to collect precise estimates of diagnostic performance. Specifically, we aimed to gather data on the sensitivity and specificity of physical findings of fever, as well as imaging techniques such as ultrasound (USG) and computed tomography (CT) scans, and the LRINEC score, in detecting Necrotizing Soft Tissue Infections (NSTI) in patients.

## METHODOLOGY

### DATA COLLECTION

For all applicable literature, a hunt was done using PubMed, Google Scholar, and Cochrane Library databases. Full-Text papers written only in English were considered. The medical subject headlines(MeSH) and keywords ‘ CT checkup for Necrotizing Soft Tissue Infections(NSTI) ’, ‘ Fever for relating NSTI ’, ‘ Hypotension for relating NSTI ’, and ‘ Diagnosis imaging for Necrotizing Soft Tissue Infections(NSTI) ’, ‘ Necrotizing Soft Tissue Infections(NSTI) ’, ‘ LRINEC score ’, ‘ USG for NSTI ’ were used. References, reviews, and meta-analyses were surveyed for fresh papers.

### INCLUSION AND EXCLUSION CRITERIA

Titles and abstracts were screened, and Duplicates and citations were removed. References of applicable papers were reviewed for possible fresh papers. Papers with detailed patient information and statically supported results were elected. The primary idea was to determine the individual accuracy of ultrasound(USG) and computed tomography(CT) scans, the LRINEC score, and clinical signs like fever and hypotension in detecting Necrotizing Soft Tissue Infections(NSTI) in cases. We included studies that compared the outgrowth and diagnostic accurateness of ultrasound(USG) and computed tomography(CT) scans, and the LRINEC score and clinical signs like fever and hypotension with surgery for suspected NSTI in the overall population involving children, Pregnant cases, and Adults. accordingly, the purpose of this study was to perform a systematic review and meta-analysis of the use of ultrasound(USG) and computed tomography(CT) reviews, and the LRINEC score and clinical signs like fever and hypotension to diagnose NSTI in the general population, ie, not restricted to one subpopulation such as pregnant cases or children. The immediate issues of interest are the sensitivity and specificity of ultrasound(USG) and computed tomography(CT) scans, the LRINEC score, and clinical signs like fever and hypotension. The inclusion criteria were as follows(1) research that handed information about the accurate diagnosis with ultrasound(USG) and computed tomography(CT) scans, and the LRINEC score and clinical sign like fever and hypotension for the diagnosis of NSTI;(2) researches published in English;(3) researches comparing of ultrasound(USG) and computed tomography(CT) scans, and the LRINEC score and clinical sign like fever and hypotension to diagnose NSTI with surgery. The exclusion criteria were(1) papers that weren’t fully manual,(2) unpublished papers, and(3) papers in other languages.

## DATA EXTRACTION

Each qualifying paper was independently estimated by two critics. Each paper was analyzed for the number of cases, age, procedure modality, and prevalence of the predecided complexities. added argumentation or discussion with the author and a third party was used to resolve conflicts. The study’s quality was assessed using the modified Jadad score. In conclusion, coinciding with PRISMA, a total of 49 RCTs with an aggregate of 11,520 cases were handpicked.

## ASSESSMENT OF STUDY QUALITY

Two authors independently assessed the quality of each contained study. This test consists of 10 questions, each with a score between 0 and 2, with 20 being the maximum possible overall score. Two authors rated each paper independently based on the below criteria. The interobserver agreement for study selection was determined using the weighted Cohen’s kappa(K) coefficient. For deciding the bias threat for RCTs, we also employed the Cochrane tool. No hypotheticals were made about any missing or unclear information. there was no sponsorship involved in collecting or examining data.

## STATISTICAL ANALYSIS

The statistical software bundles RevMan(Review Manager, version 5.3), SPSS(Statistical Package for the Social Sciences, version 20), and Excel in Stata 14 were employed to achieve the statistical analyses. The data was obtained and entered into logical software [23]Fixed- or random-effects models were applied to assess Sensitivity, Specificity, positive predictive value(PPV), diagnostic odds ratios(DOR), and relative risk(RR) with 95 percent confidence intervals to examine critical clinical issues(CIs). Diagnosis accuracy and younden index were computed for each conclusion. Individual study sensitivity and specificity were put up on Forest plots and in the receiver operating characteristic(ROC) curve. The previous odds ratio and positive and negative likelihood ratio and positive and negative post-test ratio are depicted in Fegan’s analysis.

## BIAS STUDY

The threat of bias was estimated by applying QUADAS-2 analysis. This tool includes 4 disciplines -Patient selection, Index test, Reference standard, Flow of the patients, and Timing of the Index tests. The summary of publication bias is shown in the following charts. The publication bias in patient selection was low in 15 high in 6 and unclear in 10. The index test was low in 24 and unclear in 7 papers. While the reference standard was low in 26, high in 2, and unclear in 3. The flow and timing was low in 21 and unclear in 10. The applicability concerns in patient selection was low in 27, high in 3, and unclear in 1. Reference standard was low in 28, high in 1, and unclear in 2 respectively. The index test was low in the whole 31 papers.

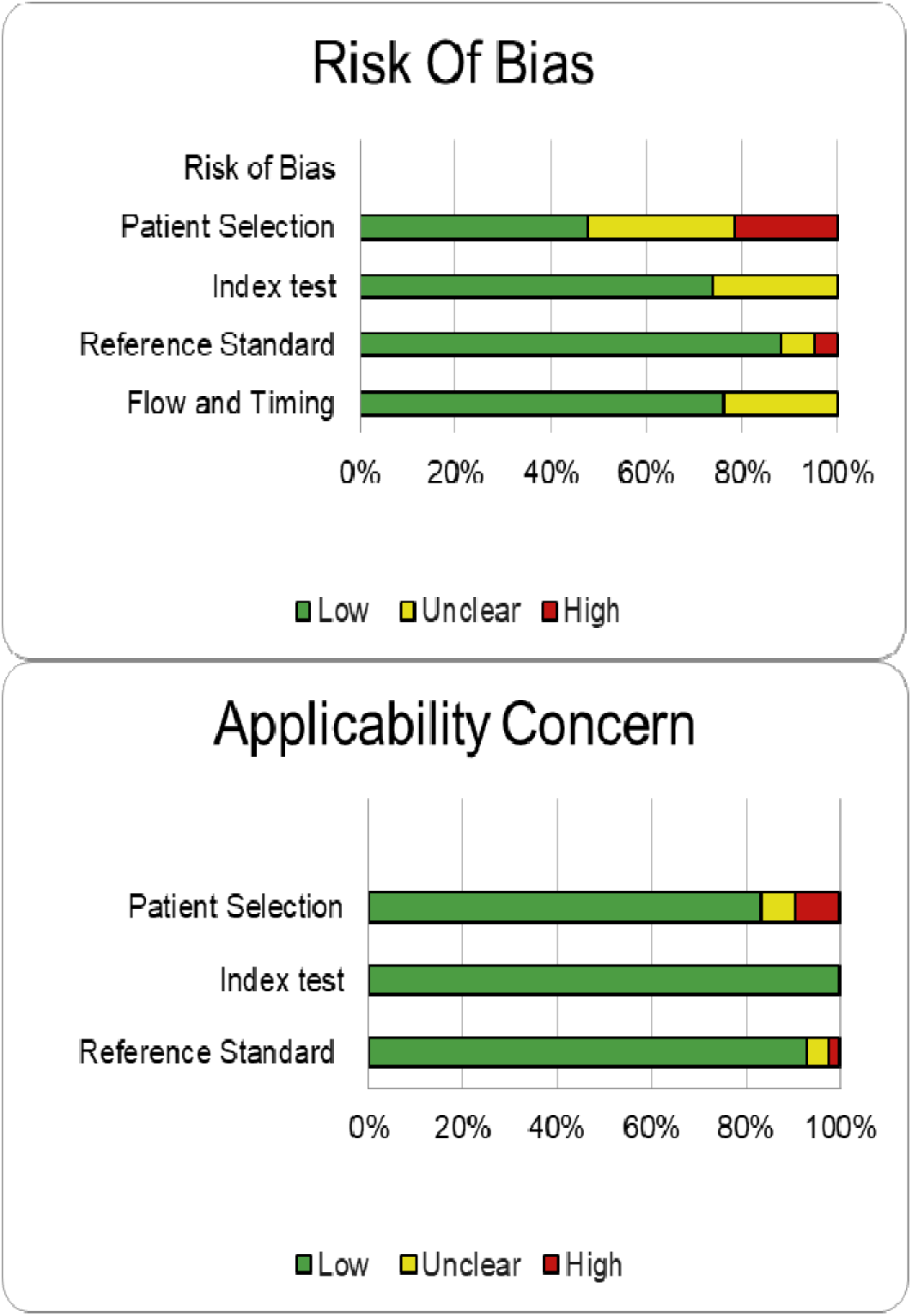

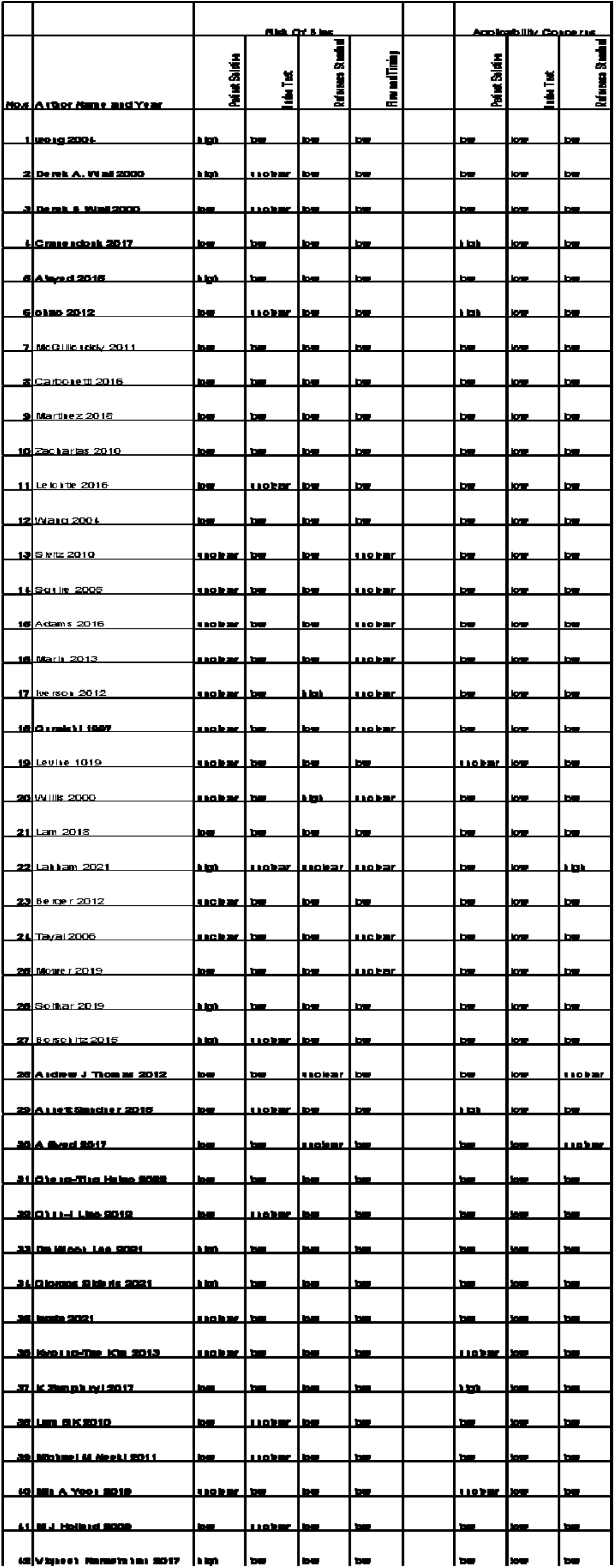

## RESULT

**Table 1:** Table of the description of papers

| Author, year | No of study participants | Mean Age | Location | Study design | Index diagnosis criteria | Reference criteria | Favoured towards |
| --- | --- | --- | --- | --- | --- | --- | --- |
| Chin-Ho Wong 2004[7] | 314 | 46-57 | Singapore | A Retrospective observational study | Hypotension, LRINEC Score $\geq 6$, fever | Surgery | LRINEC Score $\geq 6$ |
| Derek B Wall 2000 [24] | 359 | 45 | USA | A retrospective review | Hypotension | Surgery | Hypotension |
| Derek B Wall 2000[25] | 42 | 39 | USA | A retrospective review | Hypotension | Surgery | Hypotension |
| Duncan R Cranendonk 2017[26] | 54 | 51 | Netherlands | A prospective observational study | Hypotension | Surgery | Hypotension |
| Khalid Al Alayed 2015[27] | 120 | More than 50 | Canada | A retrospective review | Hypotension, fever | Surgery | Hypotension, fever |
| Wai-Nang Chao 2012[28] | 125 | 63 | Taiwan | A retrospective study | Hypotension, LRINEC Score $\geq 6$, fever | Surgery | LRINEC Score $\geq 6$ |
| Angir Soitkar 2019[29] | 166 | 47.14±15.76 | India | A prospective observational study | Fever, LRINEC Score $\geq 6$ | Surgery | LRINEC Score $\geq 6$ |
| Borschitz 2015[30] | 88 | 57.1±17.1 | Germany | A retrospective study | Fever, LRINEC Score $\geq 6$ | Surgery | Fever |
| Adam B Sivitz 2010[31] | 50 | 9.5 | USA | a prospective observational study | USG | Surgery | USG |
| Benjamin T Squire 2005[32] | 108 | 39 | USA | - | USG | Surgery | USG |
| Cynthia M Adams 2016[33] | 151 | 7 | USA | a prospective study | USG | Surgery | USG |
| Jennifer R Marin 2013[34] | 352 | 6 | USA | a prospective study | USG | Surgery | USG |
| Katrina Iverson 2012[35] | 65 | 5.2 | USA | a prospective study | USG | Surgery | USG |
| M S Quraishi 1997[36] | 23 | 2 | Ireland | A prospective study | USG | Surgery | USG |
| Marla C Levine 1019[37] | 27 | 5.3 | USA | A prospective pilot study | USG | Surgery | USG |
| Page-Willis 2000[38] | 56 | - | USA | - | USG | Surgery | USG |
| Samuel H F Lam 2018[39] | 214 | 7.5 | USA | a prospective, multicenter, cohort observational study | USG | Surgery | USG |
| Shadi Lahham 2021[40] | 95 | - | - | A Prospective study | USG | Surgery | USG |
| Tony Berger 2012[41] | 39 | - | USA | A prospective, observational study | USG | Surgery | USG |
| Vivek S Tayal 2006[42] | 126 | 42 | USA | A prospective observational Study | USG | Surgery | USG |
| William R Mower 2019[43] | 1216 | 36 | USA | A prospective observational study | USG | Surgery | USG |
| Edward A McGillicuddy 2011[44] | 305 | - | USA | A prospective study | CT | Surgery | CT |
| Francesco Carbonetti 2016[45] | 36 | - | Italy | A retrospective study | CT, LRINEC Score $\geq 6$ | Surgery | CT, LRINEC Score $\geq 6$ |
| Myriam Martinez 2018[46] | 184 | 51.4 | USA | A prospective study | CT | Surgery | CT |
| Nikos Zacharias 2010[47] | 67 | - | USA | A prospective study | CT | Surgery | CT |
| Stefan Walter<br>Leichtle<br>2016[48] | 86 | - | USA | - | CT | Surgery | CT |
| Tzong-Luen<br>Wang 2004[49] | 22 | 56-59 | Taiwan | A prospective<br>study | CT | Surgery | CT |
| Andrew J<br>Thomas<br>2012[50] | 87 | 44 | USA | A retrospective<br>review | LRINEC Score<br>≥ 6 | Surgery | LRINEC<br>Score ≥ 6 |
| Annett Sandner<br>2015[51] | 611 | - | - | - | LRINEC Score<br>≥ 6 | Surgery | LRINEC<br>Score ≥ 6 |
| A Syed<br>2017[52] | 44 | - | Malaysia | A cross-sectional<br>study involving a<br>retrospective<br>analysis | LRINEC Score<br>≥ 6 | Surgery | LRINEC<br>Score ≥ 6 |
| Cheng-Ting<br>Hsiao 2022[53] | 939 | 68 | Taiwan | A Prospective and<br>observational<br>cohort study | LRINEC Score<br>≥ 6 | Surgery | LRINEC<br>Score ≥ 6 |
| Chun-I Liao<br>2012[54] | 1627 | 60 | Taiwan | A validation<br>cohort study | LRINEC Score<br>≥ 6 | Surgery | LRINEC<br>Score ≥ 6 |
| Da Woon Lee<br>2021[55] | 201 | - | - | - | LRINEC Score<br>≥ 6 | Surgery | LRINEC<br>Score ≥ 6 |
| Giorgos Sideris<br>2021[56] | 550 | - | Greece | A retrospective<br>chart review study | LRINEC Score<br>≥ 6 | Surgery | LRINEC<br>Score ≥ 6 |
| Iwata 2021[57] | 229 | - | - | - | LRINEC Score<br>≥ 6 | Surgery | LRINEC<br>Score ≥ 6 |
| Kyoung-Tae<br>Kim 2013[58] | 30 | - | South<br>Korea | A retrospective<br>review | LRINEC Score<br>≥ 6 | Surgery | LRINEC<br>Score ≥ 6 |
| K Zemplenyi<br>2017[59] | 43 | 37-48 | USA | A Retrospective<br>Cohort Study | LRINEC Score<br>≥ 6 | Surgery | LRINEC<br>Score ≥ 6 |
| Lam SK<br>2010[60] | 285 | - | - | - | LRINEC Score<br>≥ 6 | Surgery | LRINEC<br>Score ≥ 6 |
| Michael M<br>Neeki 2011[61] | 995 | 48 | USA | A retrospective<br>chart-review study | LRINEC Score<br>≥ 6 | Surgery | LRINEC<br>Score ≥ 6 |
| Min A Yoon<br>2019[62] | 145 | 57 | South<br>Korea | A retrospective<br>study | LRINEC Score<br>≥ 6 | Surgery | LRINEC<br>Score ≥ 6 |
| M J Holland<br>2009[63] | 28 | - | Australia | A Retrospective<br>Study | LRINEC Score<br>≥ 6 | Surgery | LRINEC<br>Score ≥ 6 |
| Vignesh<br>Narasimhan<br>2017[64] | 268 | - | Australia | A Retrospective<br>Study | LRINEC Score<br>≥ 6 | Surgery | LRINEC<br>Score ≥ 6 |

### FEVER VS HISTOLOGY

A total of 5 RCTs with 813 patients were selected for the study(figure 2). Out of these tests, The value of True positive was 155, True Negative was 301, False negative was 124, and False Positive was 233. With a confidence interval of 95%, Sensitivity, specificity, and Positive Predictive values were calculated. A summary of this is available in Figure 2. The Sensitivity of the test is 0.556 with a CI of 95% in a range of (0.438 to 0.673) the mean being (0.118). The Specificity of the test is 0.564 with a CI of 95% in a range of (0.431 to 0.696) the mean being (0.133). The PPV is 0.399 with a CI of 95% in a range of (0.290 to 0.509) the mean being (0.110).

**(Figure 1).**
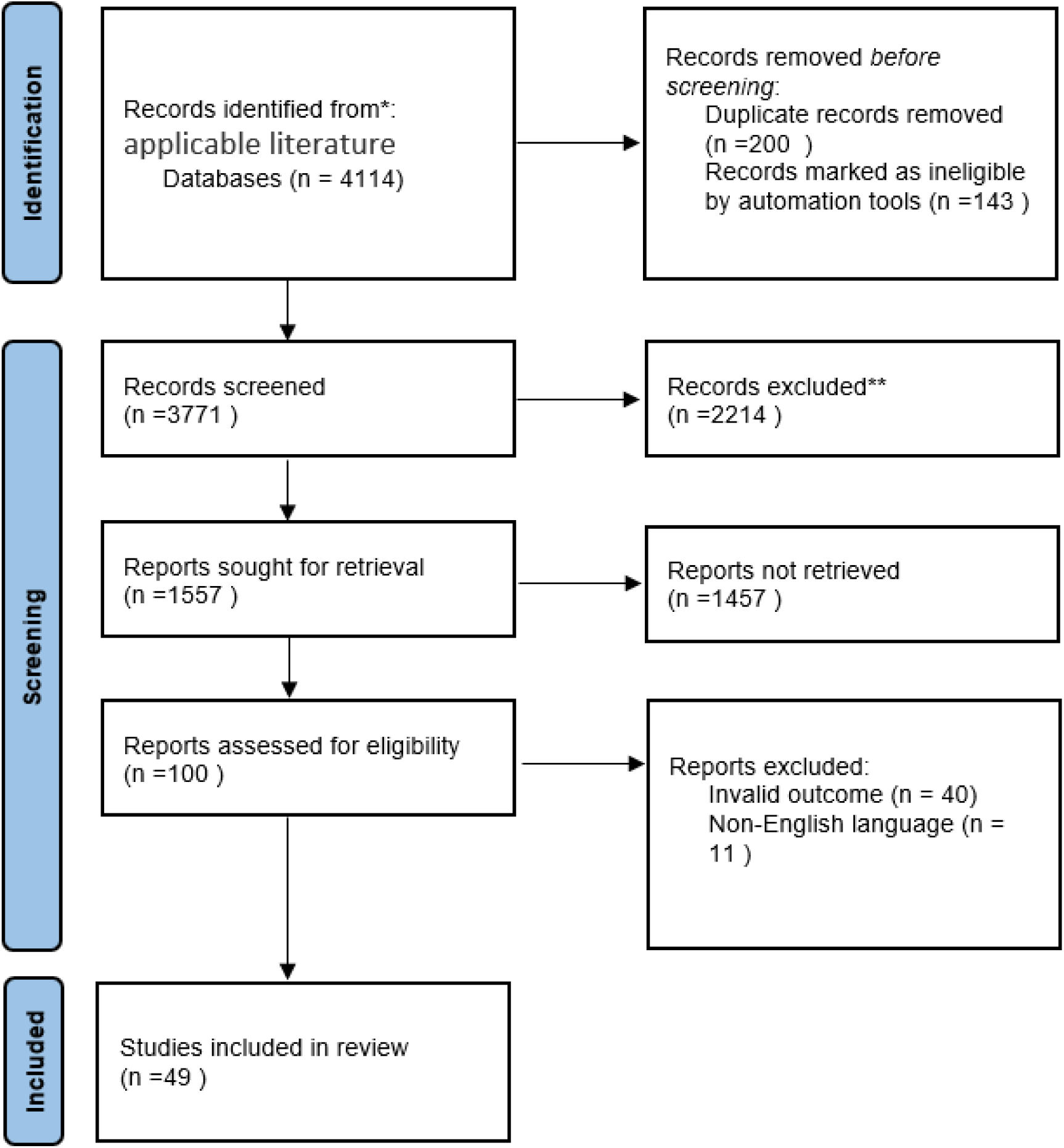
PRISMA Flowchart

**Figure 2:**
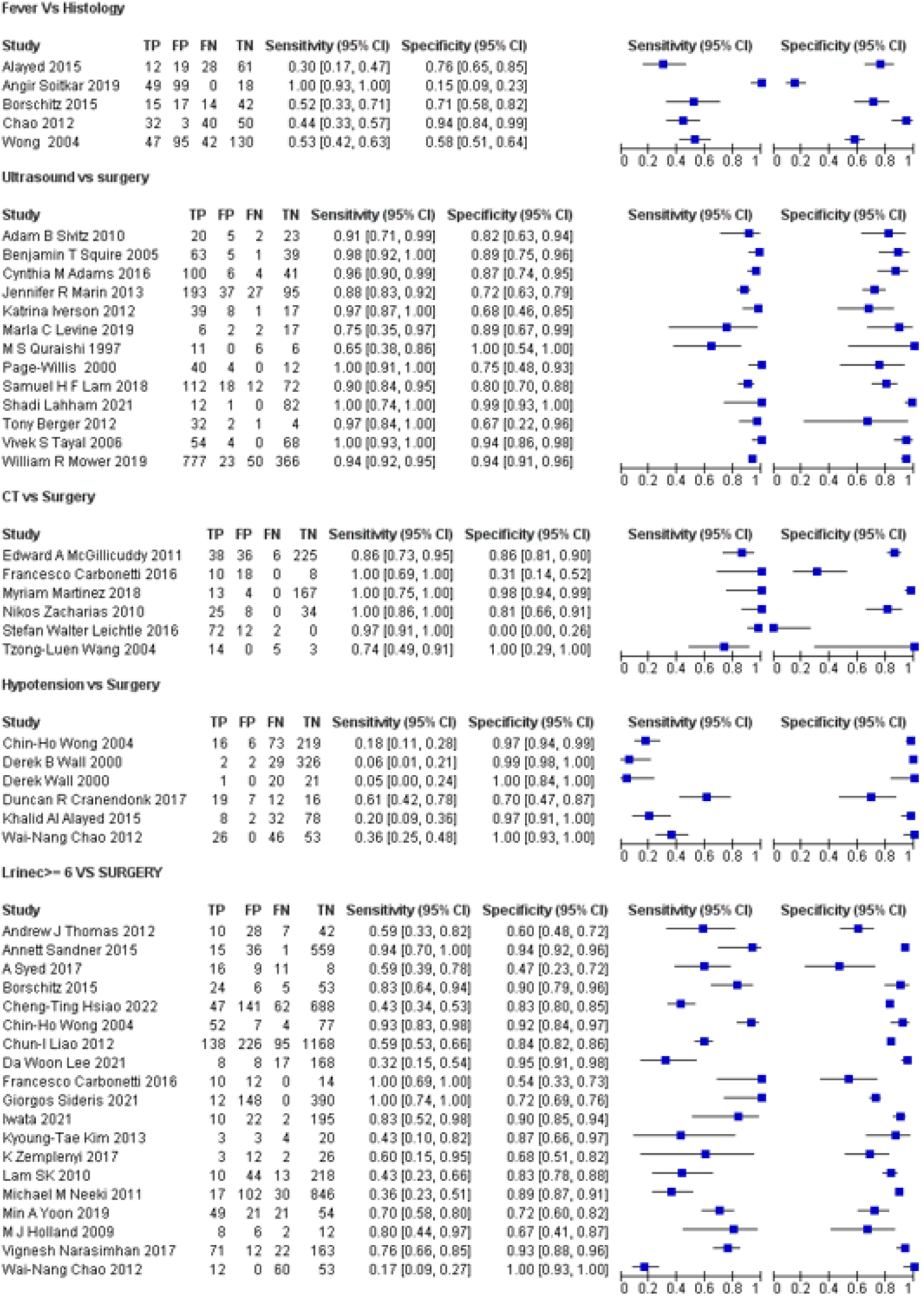
**Summary of Forest.Chart of all thepapers. Comparing the sensitivity and specificity of different studies.**

The summary of the ROC curve (Figure 3) shows that the area under the ROC (AUC) was 0.651. The overall diagnostic odds ratio (DOR) was 1.6. Diagnostic Accuracy is 0.56 and The younden Index is 0.119.

**Figure 3:**
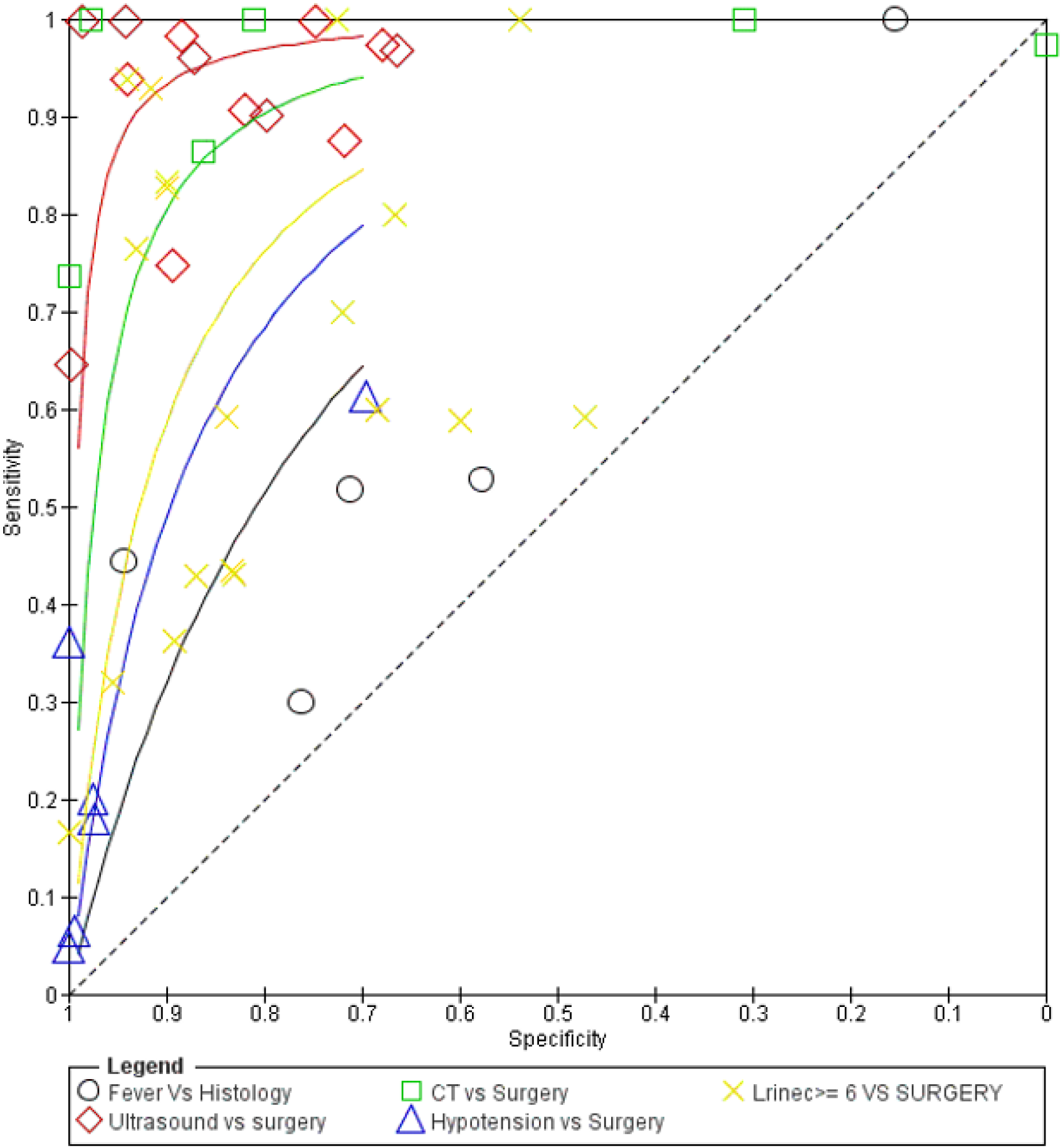
**Summary of SROC curve. Comparing The Sensitivity, Specificity, and area under the curve.**

### ULTRASOUND VS SURGERY

A total of 13 RCTs with 2522 patients were selected for the study(figure 2).Out of these tests, 7 tests showed a sensitivity of over 95%, and 2 tests provided a specificity of over 95%. And 1 test showed both specificity and sensitivity over 95%. The value of True positive was 1459, True Negative was 842, False negative was 106, and False Positive was 115. With a confidence interval of 95%, Sensitivity, specificity, and Positive Predictive values were calculated. A summary of this is available in Figure 2. The Sensitivity of the test is 0.932 with a CI of 95% in a range of (0.903 to 0.962) the mean being (0.030). The Specificity of the test is 0.879 with a CI of 95% in a range of (0.848 to 0.911) the mean being (0.031). The PPV is 0.927 with a CI of 95% in a range of (0.907 to 947) the mean being (0.020).

The summary of the ROC curve (Figure 3) shows that the area under the ROC (AUC) was 0.934. The overall diagnostic odds ratio (DOR) was 100.312. Diagnostic Accuracy is 0.912 and The younden Index is 0.812.

### CT VS SURGERY

A total of 6 RCTs with 700 patients were selected for the study(figure 2). Out of these tests, 4 tests showed a sensitivity of over 95%, and 1 test provided a specificity of over 95%. The value of True positive was 172, True Negative was 437, False negative was 13, and False Positive was 78. With a confidence interval of 95%, Sensitivity, specificity, and Positive Predictive values were calculated. A summary of this is available in Figure 2. The Sensitivity of the test is 0.930 with a CI of 95% in a range of (0.886 to 0.974) the mean being (0.044). The Specificity of the test is 0.849 with a CI of 95% in a range of (0.681 to 1.01) the mean being (0.167). The PPV is 0.688 with a CI of 95% in a range of (0.592 to 0.784) the mean being (0.096).

The summary of the ROC curve (Figure 3) shows that the area under the ROC (AUC) was 0.827. The overall diagnostic odds ratio (DOR) was 74.126. Diagnostic Accuracy is 0.870 and The younden Index is 0.778.

### HYPOTENSION VS SURGERY

A total of 6 RCTs with 1014 patients were selected for the study(figure1 2). The value of True positive was 72, True Negative was 713, False negative was 212, and False Positive was 17. With a confidence interval of 95%, Sensitivity, specificity, and Positive Predictive values were calculated. A summary of this is available in Figure 2. The Sensitivity of the test is 0.254 with a CI of 95% in a range of (0.167 to 0.340) the mean being (0.087). The Specificity of the test is 0.977 with a CI of 95% in a range of (0.928 to 1.026) the mean being (0.049). The PPV is 0.809 with a CI of 95% in a range of (0.732 to 0.886) the mean being (0.077).

The summary of the ROC curve (Figure 3) shows that the area under the ROC (AUC) was 0.620. The overall diagnostic odds ratio (DOR) was 14.244. Diagnostic Accuracy is 0.774 and The younden Index is 0.230.

### LRINEC SCORE >= 6 VS SURGERY

A total of 19 RCTs with 6471 patients were selected for the study(figure 2). Out of these tests, 2 tests showed a sensitivity of over 95%, and 2 tests provided a specificity of over 95%. The value of True positive was 515, True Negative was 4754, False negative was 358, and False Positive was 843. With a confidence interval of 95%, Sensitivity, specificity, and Positive Predictive values were calculated. A summary of this is available in Figure 2. The Sensitivity of the test is 0.590 with a CI of 95% in a range of (0.533 to 0.646) the mean being (0.056). The Specificity of the test is 0.849 with a CI of 95% in a range of (0.815 to 0.884) the mean being (0.035). The PPV is 0.379 with a CI of 95% in a range of (0.316 to 0.316) the mean being (0.063).

The summary of the ROC curve (Figure 3) shows that the area under the ROC (AUC) was 0.650. The overall diagnostic odds ratio (DOR) was 8.106. Diagnostic Accuracy is 0.814 and The younden Index is 0.439.

In figure 4 and 5, Summary of fegan’s analysis is described according to it,

**Figure 4:**
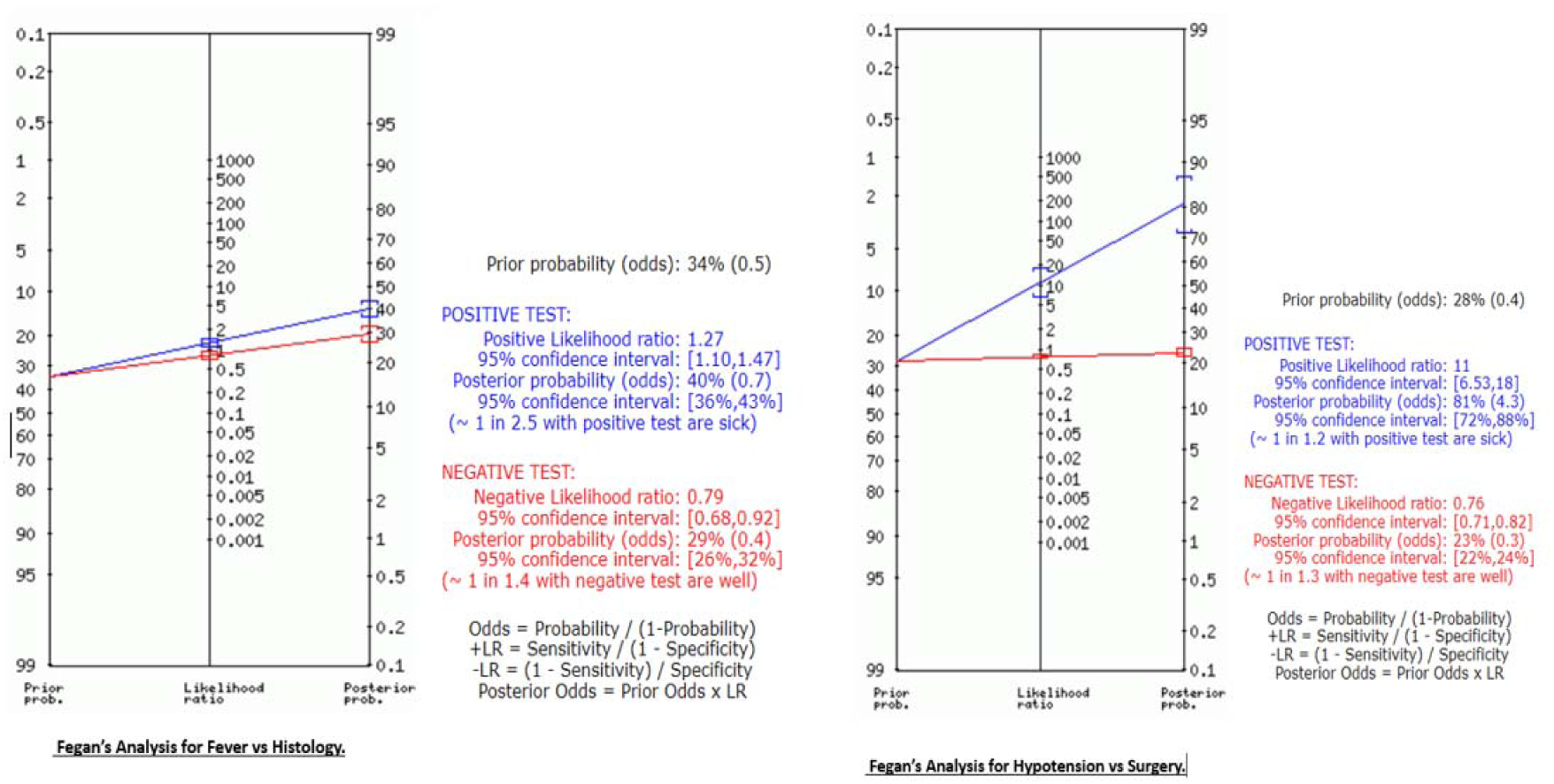
**Fegan’s Analysis of Fever and Hypotension.**

**Figure 5:**
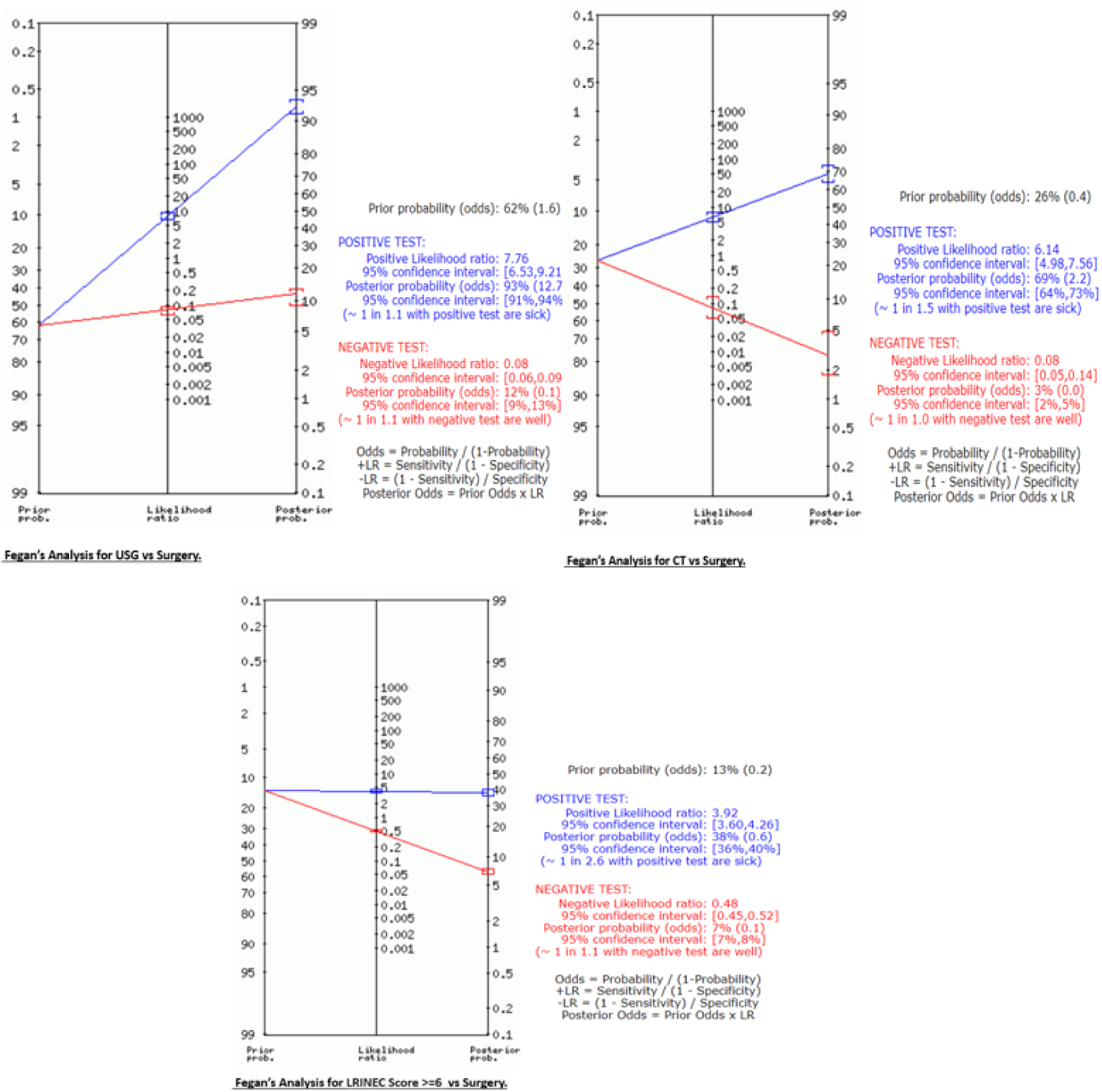
**Fegan’s Analysis of USG, CT and LRINEC Score.**

### FOR FEVER VS HISTOLOGY

The prior probability of the test was 34. The positive likelihood ratio was 1.27 and the post-test ratio was 40. The negative likelihood ratio was 0.79 and the post-test ratio was 29.

### FOR HYPOTENSION VS SURGERY

The prior probability of the test was 28. The positive likelihood ratio was 11 and the post-test ratio was 81. The negative likelihood ratio was 0.76 and the post-test ratio was 23.

### FOR USG VS SURGERY

The prior probability of the test was 62. The positive likelihood ratio was 7.76 and the post-test ratio was 93.The negative likelihood ratio was 0.08 and the post-test ratio was 12.

### FOR CT VS SURGERY

The prior probability of the test was 26. The positive likelihood ratio was 6.14 and the post-test ratio was 69.The negative likelihood ratio was 0.08 and the post-test ratio was 8.

### FOR LRINEC SCORE >= 6 VS SURGERY

The prior probability of the test was 13. The positive likelihood ratio was 3.92 and the post-test ratio was 38. The negative likelihood ratio was 0.48 and the post-test ratio was 7.

## DISCUSSION

A systematic review and meta-analysis was performed to evaluate the accuracy of physical examination findings, imaging, and LRINEC score in diagnosis of Necrotising soft tissue infection (NSTI). We gathered data from related 45 original articles constituting a total n of 928358 patients. It is essential to gain an understanding of the diagnostic accuracy of these tests to appropriately weigh the risks and benefits of using them, because the clinical implications of delayed or missed NSTI diagnosis, and that waiting for imaging or laboratory results may delay time to definitive surgical management.[1],[2]

Since there are clinical variations in presentation of NSTI, a combination of physical findings is required to make the diagnosis. But the available literature does not evaluate the various possible combination of findings to make an accurate diagnosis. And thereby, imaging modalities are used simultaneously with physical examination to increase the diagnostic accuracy of NSTI. While combining the mentioned studies, this study comprehensively compares and summarises the currently available modalities and diagnostic combinations, to figure out the most sensitive and specific diagnostic tool for NSTI. The description of NSTI and clinical diagnosis, based on patient risk factors and findings on physical examination as mentioned in the literature [2]does not have very high sensitivity clinically.

Several specific and pathognomic findings[1] for diagnosis of NSTI are available in the literature, out of which we are assessing fever and hypotension for this review. We found that both fever and hypotension have low sensitivity for diagnosis of NSTI, therefore the absence of either of the finding would not rule out the disease. This becomes clearly evident on plotting the data gathered from the articles to measure the diagnostic accuracy of fever vs histology for NSTI. As seen in the Table A (Fever table), the sensitivity (55.6%), specificity (56.4%) and AUC (0.651) being low, it cannot be reliably utilised as a sole diagnostic parameter. A low PPV of 39% again makes it an unreliable parameter. Patients with suspected NSTI will still have to undergo further testing and confirmation before finally proceeding with the definitive surgical management [4]

Similarly, the sensitivity (25.4%) for hypotension (Hypotension table) as a diagnostic parameter is significantly low as well, when compared to imaging modalities. Even though the specificity (97.7%) and PPV (80.9%) of hypotension is high enough to diagnose NSTI, there has been evidence of appearance of hypotension and shock in the more advanced disease [3]Hence, relying on this parameter as a sole indicator can potentially lead to delay in diagnosis and immediate essential surgical management.

On assessing for the use of imaging modality to diagnose NSTI, we analysed the accuracy of CT scan and point-of-care USG. Studies with pragmatically broadened CT criteria to assess the signs of NSTI on the scan, the overall sensitivity calculated was as high as 93%, with sensitivity being 84.9% (CT Table). High PPV (68.8%) and AUC (0.827) also makes it quite a reliable imaging method for diagnosing and confirming NSTI. However, even though CT has a high specificity, it has several components and findings that can have a wide range of diagnostic possibilities, similar to physical examination findings. Which may necessitate need for a standard universal reporting score or checklists for CT in suspected cases of NSTI. Apart from that, another major factor is limited availability of CT imaging. Also, CT scan imaging is time consuming which potentially leads to delay in definitive surgical management. Taking all these factors into account, even though CT scan has a relatively strong accuracy in diagnosing NSTI, delay in surgical consultation and management should not occur.

Point-of-care ultrasound, a widely available bedside modality has been studied broadly in literature and has a significant role in diagnosis of NSTI in current time. On assessing and combining data from the previous studies done on role of point-of-care USG in diagnosing NSTI, data from a bigger pool of patient population can be analysed, thereby increasing the power of the analysis made (USG Table). The overall sensitivity of 93.2%, with high specificity (87.9%), narrow CI and AUC (0.934) makes it a fairly accurate diagnostic test for NSTI compared to those mentioned here. The easy availability of this modality, high diagnostic accuracy and its ability to be used without significant delay of surgical consultation makes it a highly preferred test to confirm the diagnosis of NSTI in clinical setting.

We also analysed the diagnostic accuracy of LRINEC score [7] which is a widely used diagnostic modality in practice in current times [1], [2] The LRINEC score of >=6, which is clinically considered to have “moderate” risk of NSTI was calculated (LRINEC Table) to be poorly sensitive (59%) and only moderately specific (84.9%) with AUC of 0.650. Since computation and calculation of LRINEC score requires laboratory tests, a delay in surgical consultation may take place leading to worse outcomes [8]. The overall LRINEC scoring has limitations and less than desired accuracy to be used for clinical practice.

This review was performed using comprehensive search and review of the existing literature. Misdiagnosis remains a significant issue for NSTI, especially in populations considered to be at moderate to high risk, i.e., in the populations with higher prevalence (e.g.: diabetics) [9]. A subgroup analysis for a high-risk group with a higher pre-test probability has not been done, which if done might reveal increase in accuracy of the modality being analysed. The inadequate literature on the accuracy of the diagnostic tests when compared between the high-risk cohort to that of a low-risk cohort, and the inconclusive details necessitates further investigations in the subgroups accordingly. Further limitations include the heterogeneity and quality of the studies included in this metanalysis. None of the studies included here have evaluated the diagnostic accuracy of NSTI based on the body site or the total body surface area which are important prognostic factors, which when taken into consideration may alter the accuracy of the said diagnostic parameter. Finally, in addition to varied study designs of the studies included here, several studies do not mention the nature of blinding and therefore the potential bias in the interpretation of the diagnostic tests.

## CONCLUSION

In our systematic review and analysis, we can conclude that relying on individual physical examination signs such as fever or hypotension was not a reliable method for diagnosing NSTI. Even though CT scans were found to have superior sensitivity and specificity, their availability may be limited in some centers and they may not be appropriate for unstable patients. Additionally, our findings indicate that the LRINEC score is not a reliable indicator of NSTI diagnosis, as a low score does not necessarily rule out the possibility of NSTI. POCUS is a relatively accurate diagnostic test for NSTI when compared to other methods. Its easy availability, high accuracy, and ability to be used without delaying surgical consultation make it a preferred diagnostic tool for confirming NSTI in clinical practice.

## Data Availability

All data produced in the present work are contained in the manuscript

